# A Systematic Review and Meta-Analysis on the Effectiveness of Bivalent mRNA Booster Vaccines against Omicron Variants

**DOI:** 10.1101/2024.01.10.24301107

**Authors:** Shangchen Song, Zachary J. Madewell, Mingjin Liu, Yu Miao, Shaolin Xiang, Yanan Huo, Shoumi Sarkar, Amily Chowdhury, Ira M. Longini, Yang Yang

**Affiliations:** Department of Biostatistics, College of Public Health and health Professions, University of Florida, Gainesville, FL, USA; Department of Statistics, Franklin College of Arts and Sciences, University of Georgia, Athens, GA, USA; Gilead Sciences, Inc, Foster City, CA, USA; Department of Computer Science, Franklin College of Arts and Sciences, University of Georgia, Athens, GA, USA

**Keywords:** Bivalent, Omicron, Vaccine Effectiveness, Meta-Analysis, Infection, Severe disease

## Abstract

**Background:** A global shift to bivalent mRNA vaccines is ongoing to counterbalance diminishing monovalent vaccine effectiveness (VE) due to the evolution of SARS-CoV-2 variants, yet substantial variation in the bivalent VE exists across studies and a complete picture is lacking.

**Methods:** We searched papers evaluating SARS-CoV-2 bivalent mRNA vaccines on PubMed, Web of Science, Cochrane Library, Google Scholar, Embase, Scopus, bioRxiv, and medRxiv published from September 1st, 2022, to November 8th, 2023. Pooled VE against Omicron-associated infection and severe events was estimated in reference to unvaccinated, ≥2 monovalent doses, and ≥3 monovalent doses.

**Results:** From 630 citations identified, 28 studies were included, involving 55,393,303 individuals. Bivalent boosters demonstrated superior protection against symptomatic or any infection compared to unvaccinated, ≥2 monovalent doses, and ≥3 monovalent doses, with corresponding relative VE estimated as 53.5% (95% CI: - 22.2-82.3%), 30.8% (95% CI: 22.5-38.2%), and 28.4% (95% CI: 10.2-42.9%) for all ages, and 22.5% (95% CI: 16.8-39.8%), 31.4% (95% CI: 27.7-35.0%), and 30.6% (95% CI: -13.2-57.5%) for adults ≥60 years old. Pooled bivalent VE estimates against severe events were higher, 72.9% (95% CI: 60.5-82.4%), 57.6% (95% CI: 42.4-68.8%), and 62.1% (95% CI: 54.6-68.3%) for all ages, and 72.0% (95% CI: 51.4-83.9%), 63.4% (95% CI: 41.0-77.3%), and 60.7% (95% CI: 52.4-67.6%) for adults ≥60 years old, compared to unvaccinated, ≥2 monovalent doses, and ≥3 monovalent doses, respectively.

**Conclusions:** Bivalent boosters demonstrated higher VE against severe outcomes than monovalent boosters across age groups, highlighting the critical need for improving vaccine coverage, especially among the vulnerable older subpopulation.

## Introduction

The emergence of the SARS-CoV-2 Omicron (B.1.1.529) variant in late 2022 significantly undermined previous control measures in the continuing COVID-19 pandemic. Despite significant progress in vaccination efforts worldwide, Omicron continues to pose a substantial threat, causing a significant number of severe COVID-19 cases and fatalities due to continuously evolving immune escape. Monovalent mRNA vaccines, initially designed for ancestral SARS-CoV-2 variants, demonstrated high effectiveness in preventing severe COVID-19 caused by the Omicron variants [1,2]. One or two monovalent mRNA booster doses also reduced hospitalizations and fatalities [3]. However, their effectiveness waned over time, particularly against Omicron [4,5]. The Omicron sub-lineages from BA.1 to BA.5 and subsequent variants such as XBB.1.5, EG.5, and FL.1.5.1 showcased the virus’s adaptability through genetic mutations in the spike protein, making it increasingly distinct from the original wild type [6]. Laboratory studies demonstrated that antibodies were less effective at neutralizing these emerging subvariants compared to ancestral variants [7,8].

In response to reduced effectiveness of monovalent boosters, bivalent mRNA booster vaccines containing spike sequences from the original SARS-CoV-2 strain and Omicron subvariants (BA.1 or BA.4-5) emerged as a potential strategy to enhance protection against severe clinical outcomes. Laboratory studies demonstrated that bivalent vaccines significantly increased neutralizing activity against subvariants like BA.4, BA.5, BA.2.75, BQ.1.1, and XBB compared to the monovalent vaccines [9,10]. On September 1, 2022, the Advisory Committee on Immunization Practices (ACIP) recommended these vaccines to address the reduced effectiveness of monovalent vaccines [11]. The U.S. Food and Drug Administration (FDA) subsequently approved Moderna and Pfizer-BioNTech bivalent formulations as single booster doses for individuals who completed primary vaccination series or received monovalent boosters [12]. Since September 2022, many countries have transitioned from monovalent boosters to bivalent boosters.

Recent studies increasingly demonstrate the higher effectiveness for bivalent boosters compared to monovalent vaccines against infection and severe disease [13–15]. Meta-analyses provide an integrative assessment of the effectiveness of bivalent boosters by aggregating data from multiple studies, enhancing statistical robustness and generalizability, while controlling for differences between individual studies. One review and case study reported vaccine effectiveness (VE) for bivalent boosters against hospitalization and infection, but was limited to a few early studies [16]. Herein, we synthesize contemporary literature to report absolute and relative VE of bivalent boosters against infection and severe COVID-19 compared to no vaccination, two or more monovalent doses, and three or more monovalent doses, respectively.

## Methods

This analysis followed the Preferred Reporting Items for Systematic Reviews and Meta-analyses (PRISMA) reporting guidelines.

### Data Sources and Searches

We systematically searched databases and platforms including PubMed, Web of Science, Cochrane Library, Google Scholar, Embase, Scopus, and preprint servers (bioRxiv and medRxiv) for papers published between September 1st, 2022 and November 8th, 2023. We applied Boolean combinations of the following keywords to identify relevant publications: “SARS-CoV-2”, “COVID-19”, “2019nCoV”, “bivalent booster”, “effectiveness”, “efficacy”, “test-negative”, “case-control”, “cohort study”, “Omicron”, “infection”, “infected”, “hospitalization”, “hospital admission”. Details about the search procedures are available in Supplementary material A.1. Publication language was not restricted, and reference lists of selected papers were also screened for additional studies.

### Study Selection

Studies were chosen based on the PICOS (Participant, Intervention, Comparator, Outcome, Study Type) criteria [17] as detailed in Supplementary Material A.2. We focused on original research using observational studies, e.g., test-negative design study or cohort study, that reported estimates for the effectiveness of COVID-19 bivalent boosters against Omicron-related infections or severe events in reference to either no vaccination or monovalent vaccine doses. The COVID-19 bivalent boosters include BA.1 type and BA.4/BA.5 type mRNA vaccines manufactured by Pfizer-BioNTech and Moderna. We excluded studies that (i) did not assess VE (e.g., evaluated neutralizing antibodies); (ii) targeted special populations (e.g., patients with kidney disease); (iii) focused on relative VE between different types or doses of bivalent vaccines; (iv) reported VE results for a mixture of bivalent and monovalent vaccines (unless bivalent representation was 90% or higher); or (v) examined outcomes unrelated to COVID-19. Our analysis included all available age groups, and we did not seek additional data from authors.

After duplicates were removed, studies were initially sifted based on titles and abstracts by three independent teams, each with three researchers. Full texts of potential matches were then independently reviewed by the same three teams. Any disagreements were discussed until a consensus was reached. Preprints were checked and updated with their most recently published version if available as of January 5th, 2024. Zotero was used for literature management.

### Data Extraction and Quality Assessment

The following information was independently extracted from the included studies: title, authors, publishing journal and year, study region, duration and design, statistical method, definition of VE, circulating subvariants, types and doses of bivalent and reference vaccines, time from vaccination to testing, age group, adjusted VE point estimate and 95% confidence intervals, and confounders adjusted for. When available, the raw numbers of vaccinated and unvaccinated individuals among cases and controls were also recorded.

Study quality and risk of bias of each study were independently assessed by three researchers using the Newcastle-Ottawa Scale (NOS). Studies were assigned up to 9 points according to participant selection (4 points), study comparability (1 point), and outcome of interest (4 points). A score >7 was considered as high quality, 5–6 as medium, and <5 as low, and studies classified as low quality were excluded from the meta-analysis. Publication bias was also evaluated by Egger’s test, Begg and Mazumdar rank correlation, and funnel plots when at least ten studies were available, with statistical significance achieved if p-value < 0.1. Upon detection of publication bias, we used the Duval and Tweedie trim-and-fill method [18] to adjust the analysis, where missing effect sizes were imputed to achieve symmetry.

### Data Synthesis and Analysis

We grouped bivalent VEs into three categories based on the reference arm: (1) unvaccinated, (2) two doses of monovalent vaccines, and (3) three or more doses of monovalent vaccines. This categorization aimed to assess VE variations across different previous vaccination benchmarks. Notably, due to the recent introduction of bivalent vaccines, most VE estimates from the studies we gathered pertain to short-term effects, typically within 120 days. If a study reported VE estimates for more granular time intervals than desired, we combined these estimates using an inverse variance weighted (IVW) averaging method to synthesize a VE estimate for the desired time interval. For example, if a study reported VE estimates for age groups 60–69 years, 70–79 years and ≥80 years old, we synthesize the three VE estimates into a single measure for individuals aged ≥60 years old. We evaluated VE for the entire vaccine-eligible population, pooling all age groups, and specifically for senior adults aged 60 and above. We assessed VE against the Omicron subvariants for both infections and severe outcomes. We did not distinguish between any infection and symptomatic infection, even though some studies focused on the latter; the implications of this approach are discussed. Severe outcomes are defined as hospitalization or death. We did not differentiate between the two types/manufacturers of bivalent vaccines as most studies reported combined VE for these vaccines.

To increase sample size, we pooled VE estimates from both test-negative studies and cohort studies. Test-negative studies used either conditional logistic regression or multivariate logistic regression with calendar time adjustment, whereas cohort studies mostly used Cox regression. Notably, simulation results in a previous study showed that logistic regression adjusting for calendar time closely approximated conditional logistic regression matched on calendar time, and the latter shares the same form as the partial likelihood of Cox regression [19]. In cases where studies did not directly report VE but provided Odds Ratios (OR) or Hazard Ratios (HR), VE was derived using the formulas: (1 – OR) × 100% or (1 – HR) × 100%, respectively. We computed pooled VE estimates and their corresponding 95% confidence intervals using a random-effect meta-analysis with restricted maximum likelihood estimation. To assess between-study heterogeneity, we employed the 1^2^ statistic, with thresholds of 25%, 50%, and 75% denoting low, moderate, and high heterogeneity, respectively. All statistical procedures and visual representations were conducted using the *metafor* R package (version 4.0.5) [20].

## Results

### Study selection and characteristics

We obtained 630 articles from all searched databases (18 from PubMed, 6 from Web of Science, 29 from Embase, 425 from Scopus, 2 from Cochrane Library, 46 from medRxiv, 4 from bioRxiv, and 100 from Google Scholar). After removing duplicates, 539 articles remained, of which 87 passed the screening of title, abstracts and keywords and underwent full text review. After further quality control such as removing duplicates and special populations, of these 87 articles as well as 2 additional articles identified through reference lists of eligible articles, 28 articles [13–15,21–45] were formally included in this meta-analysis (Figure 1).

**Figure 1.**
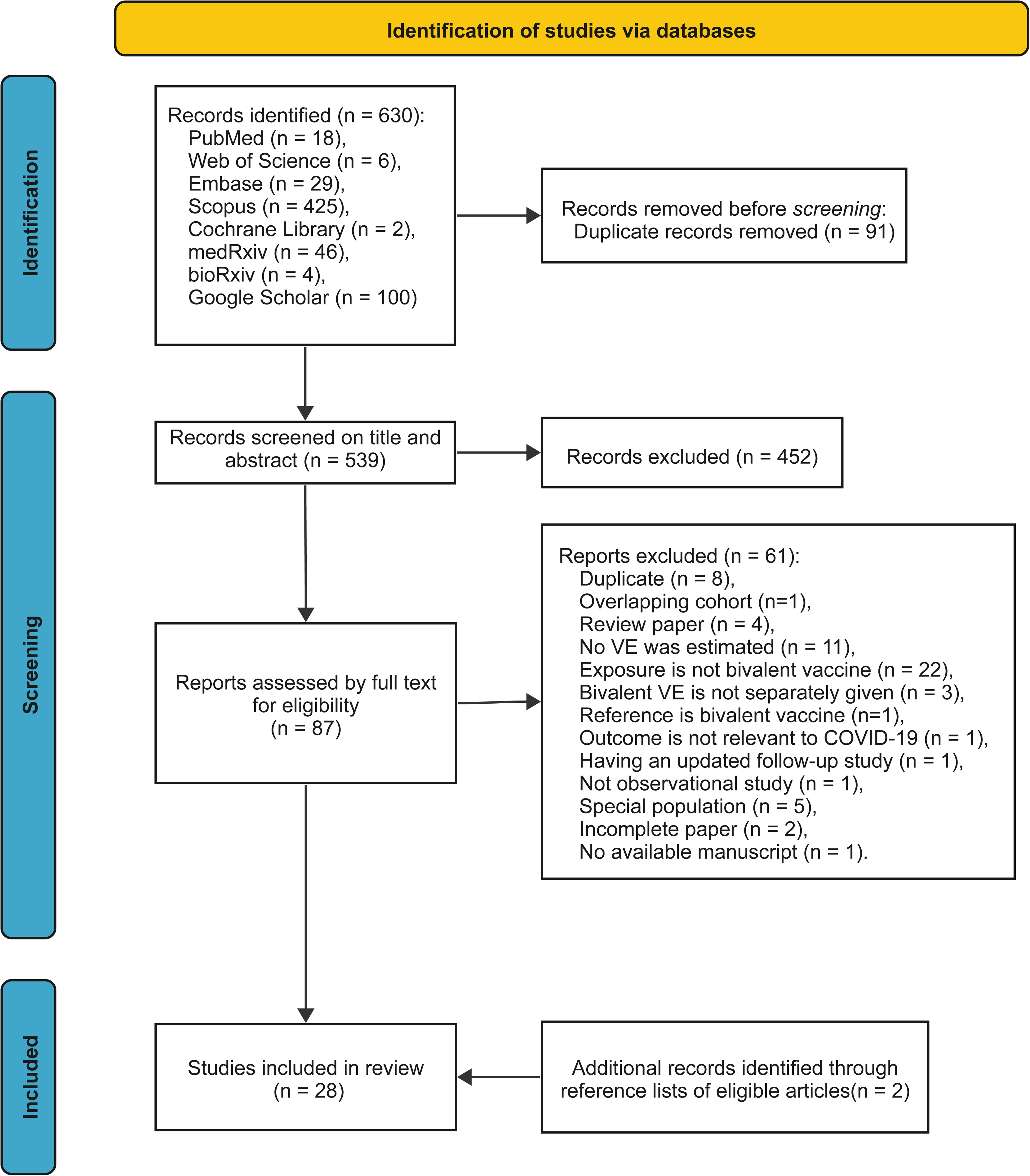
Summary of evidence search and selection.

Among the 28 papers, nine studies were conducted in the U.S., three in the U.K., three in Italy, two in each of Canada, Japan, Korea, one in each of France, Israel, Netherlands, Norway, Qatar and Singapore, and a multi-national study conducted across Denmark, Finland, Norway, and Sweden. There were 12 test-negative design studies and 16 cohort studies. Regarding the Omicron subvariants in circulation, 20, 13, and 16 studies involved BA.4/5, BQ, and XBB lineages, respectively. Most studies reported multiple VE estimates, stratified by the type of vaccine or the type of outcome. In total, there were 456 VE estimates, including 219 for the BA.1 bivalent booster, 191 for BA.4/5 bivalent booster, and 46 for mixed cohorts receiving BA.1 or BA.4/5 bivalent boosters. By the type of outcome, 36, 64, and 356 VE estimates were reported for symptomatic infection, any infection, and severe events, respectively. After screening and synthesizing VE estimates, 20 were available for calculating the pooled VE against symptomatic or any infection and 31 for the pooled VE against severe outcomes. In all included studies, the number of doses of bivalent boosters are greater or equal to the number of doses of the reference vaccination regimens, which included unvaccinated, monovalent full course (two doses of monovalent vaccines), and monovalent boosters (3–4 doses of monovalent vaccines).

### Vaccine effectiveness against Omicron symptomatic infection or any infection

The VE estimates for the bivalent boosters against Omicron symptomatic or any infection were summarized for all age groups combined in Figure 2 and for seniors aged 60 years or older in Figure 3. Only two studies reported a bivalent booster VE with unvaccinated as the reference, with a large inter-study difference, and the combined VE was 53.5% (95% CI: -22.2–82.3%) for adults aged 16 or over (Fig. 2). When the reference was monovalent full course (2 monovalent doses), the pooled bivalent VE from seven studies was 30.8% (95% CI: 22.5–38.2%) for all ages. When the reference was monovalent boosters (3 or more monovalent doses), the pooled bivalent VE from seven studies was estimated to be 41.8% (95% CI: 9.0–62.8%) for all adults ≥18 years (Fig. 2). The large uncertainty of this VE estimate came from the study of Tan et al. (2023) in Singapore which reported a VE against symptomatic infection of 83.0% with a tight 95% CI of 82.0–84.0%, much higher than the other studies (1^2^ statistic=99%). After excluding this study, the pooled VE for all ages was 28.4% (95% CI: 10.2–42.9%). For seniors over 60, one study in the U.S., Rudolph, et al. (2023), reported bivalent VE estimates against the unvaccinated and those with ≥2 monovalent doses as 22.5% (95% CI: 16.8–39.8%) and 31.4% (95% CI: 27.7–35.0%), respectively (Fig. 3). When considering ≥3 monovalent doses as a reference, the pooled VE from four studies was 49.0% (95% CI: -2.1–74.5%), but excluding Tan et al. (2023), it was 30.6% (95% CI: -13.2–57.5%). Combining all three reference vaccination groups, the pooled overall VE against symptomatic or any infection was estimated to be 38.7% (95% CI: 23.6–50.8%) for all ages and 42.5% (95% CI: 8.6–63.9%) for adults aged 60 years or older or, if excluding the Singapore study, 32.4% (95% CI: 22.6–41.0%) for all ages and 29.3% (95% CI: 7.2–46.1%, 5 studies) for seniors (Fig. 3).

**Figure 2.**
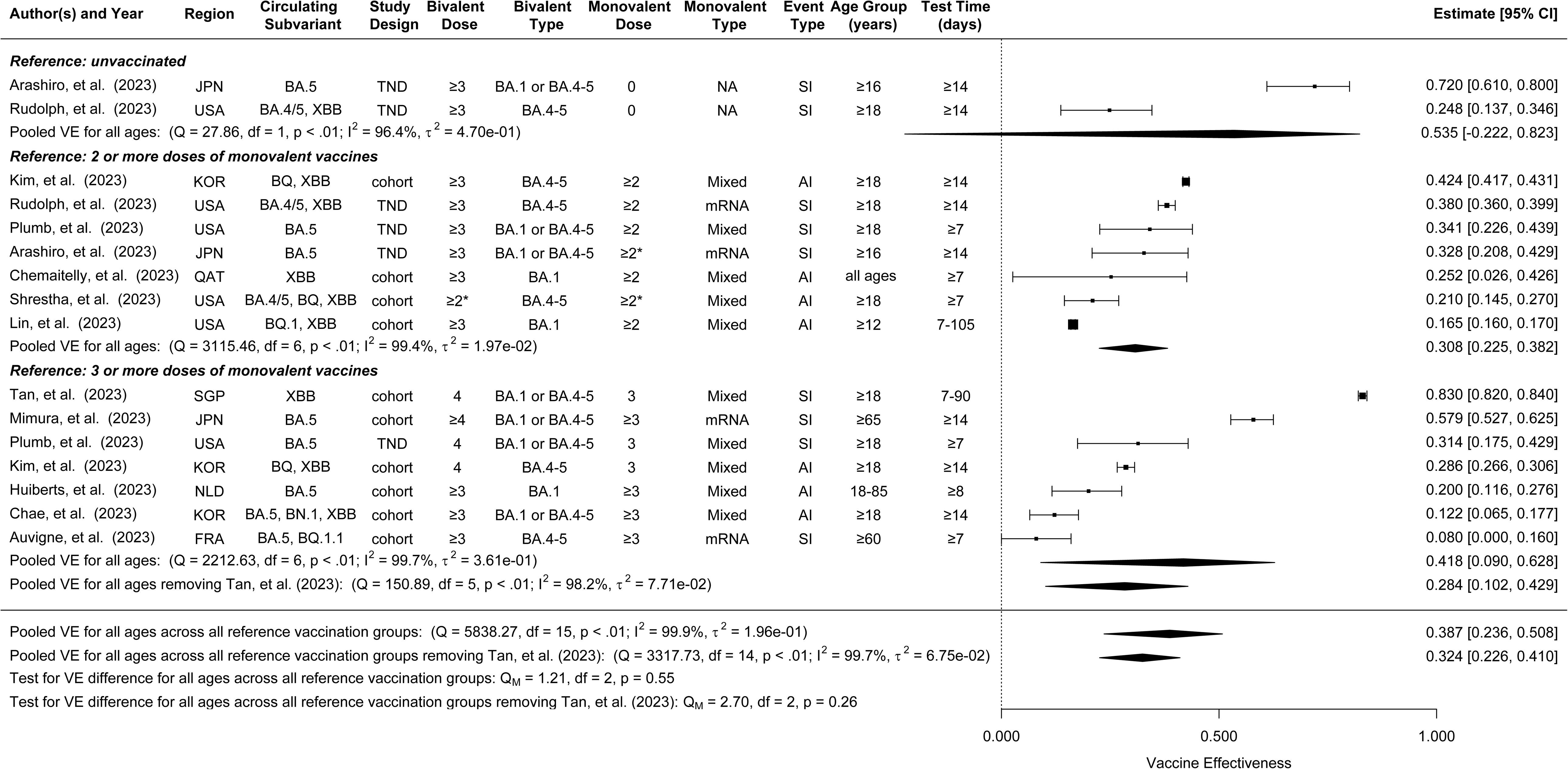
Forest plot for relative effectiveness of bivalent vaccines compared to different reference vaccine groups against any infection or symptomatic infection for all ages. Statistics Cochran’s Q, I^2^ and T^2^ measure the heterogeneity between studies. End points of the studies are either symptomatic infection (SI) or any infection (AI). TND: test-negative design study. cohort: cohort study. NA: not applicable. Mixed: containing non-mRNA and mRNA monovalent vaccines. ≥2*: the majority (≥80%) received 2 or more doses.

**Figure 3.**
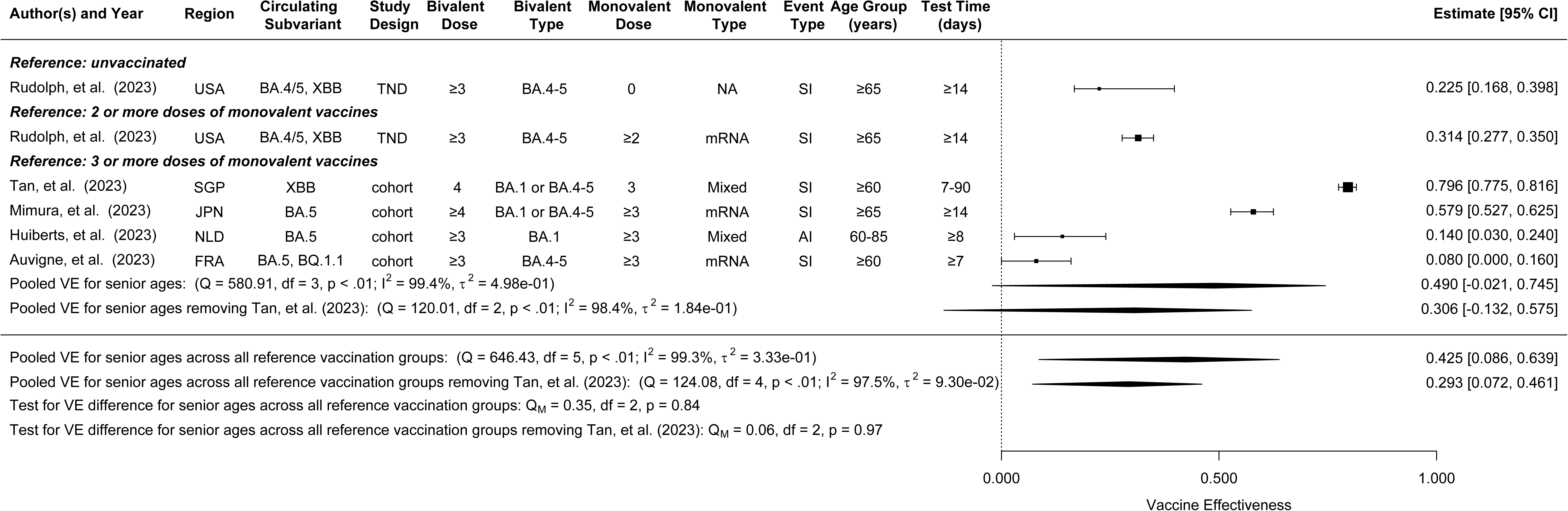
Forest plot for relative effectiveness of bivalent vaccines compared to different reference vaccine groups against any infection or symptomatic infection for senior adults ≥ 60 years old. Statistics Cochran’s Q, I^2^ and T^2^ measure the heterogeneity between studies. End points of the studies are either symptomatic infection (SI) or any infection (AI). TND: test-negative design study. cohort: cohort study. NA: not applicable. Mixed: containing non-mRNA and mRNA monovalent vaccines.

### Vaccine Effectiveness against Omicron-associated Severe Events

The pooled bivalent VE estimates against Omicron-associated severe events for all ages were 72.9% (95% CI: 60.5–82.4%, 7 studies), 57.6% (95% CI: 42.4–68.8%, 8 studies), and 68.5% (95% CI: 56.4–77.2%, 10 studies) when the reference group was unvaccinated, ≥2 monovalent doses, and ≥3 monovalent doses, respectively (Fig. 4). In the third category, the pooled VE excluding Tan et al. (2023), which also reported a higher VE estimate against severe events than the rest studies, was 62.1% (95% CI: 54.6–68.3%, 9 studies). Combining the three reference groups, the overall pooled VE against severe events was estimated to be 66.5% (95% CI: 59.2–72.6%, 25 studies) for all ages, or 64.7% (95% CI: 57.7–70.6%, 24 studies) when excluding Tan et al. (2023). For those aged 60 or over, the pooled bivalent VE estimates were 72.0% (95% CI: 51.4– 83.9%, 4 studies), 63.4% (95% CI: 41.0–77.3%, 3 studies), and 60.7% (95% CI: 52.4–67.6%, 8 studies) when the reference group was unvaccinated, ≥2 monovalent doses, and ≥3 monovalent doses, respectively (Fig. 5). The pooled bivalent VE estimates combining all reference groups was 65.2% (95% CI: 57.4–71.5%, 15 studies) for senior adults.

**Figure 4.**
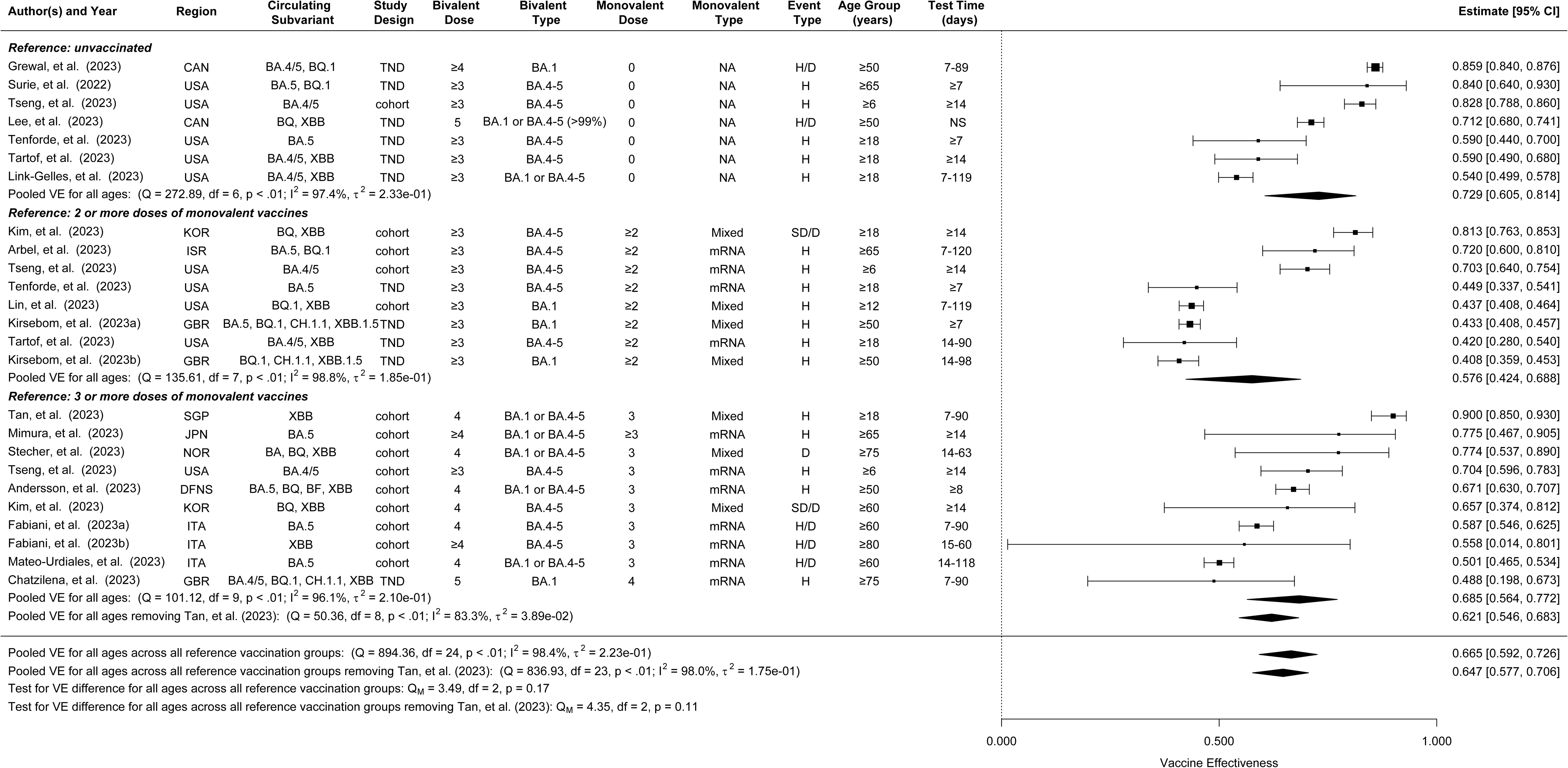
Forest plot for relative effectiveness of bivalent vaccines compared to different reference vaccine groups against severe outcomes for all ages. Statistics Cochran’s Q, I2 and τ^2 measure the heterogeneity between studies. End points of the studies are hospitalization (H), Death (D), hospitalization or death (H/D), severe disease or death (SD/D). TND: test-negative design study. cohort: cohort study. NA: not applicable. NS: not specified. Mixed: containing non-mRNA and mRNA monovalent vaccines.

**Figure 5.**
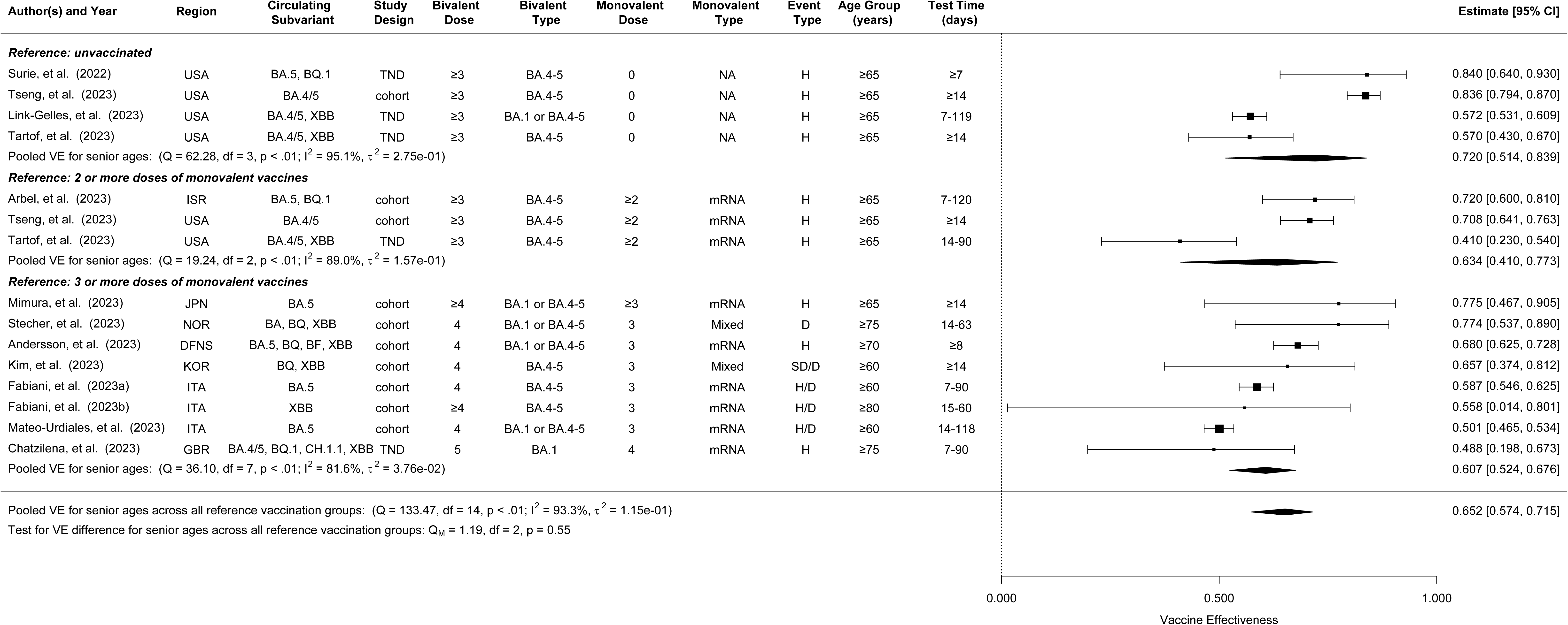
Forest plot for relative effectiveness of bivalent vaccines compared to different reference vaccine groups against severe outcomes for senior adults ≥60 years old. Statistics Cochran’s Q, I2 and τ^2 measure the heterogeneity between studies. End points of the studies are hospitalization (H), Death (D), hospitalization or death (H/D), severe disease or death (SD/D). TND: test-negative design study. cohort: cohort study. NA: not applicable. Mixed: containing non-mRNA and mRNA monovalent vaccines.

### Assessment of Publication Bias

The only detected publication bias was in the pooled estimate of the overall VE of the bivalent boosters against severe events in all age groups (Egger’s test p = 0.097, Begg’s test p = 0.236) as shown in Figure 6. Results were corrected for these biases using the trim-and-fill method.

## Discussion

In this systematic review and meta-analysis, we synthesized the available evidence from 28 studies, involving 55,393,303 individuals. We found that the bivalent COVID-19 booster doses (vaccinated as the third or subsequent doses) in addition to the previous COVID-19 vaccine series provided about 30-50% VE against infection or symptomatic infection with Omicron subvariants, in reference to unvaccinated or monovalent dose regimens. The bivalent VE against severe clinical outcomes appeared to be more robust, reaching 60-70% for various reference groups. These results underscore the importance of swift adaption of vaccine development to the evolutional path of the SARS-CoV-2 family. These real-world findings resonate with early immunogenicity studies showing higher antibody responses induced by the bivalent vaccines compared to the monovalent vaccines [46–48].

A notable observation from our analysis is that the bivalent vaccine efficacies among individuals aged ≥60 years were similar to those among the general population. For symptomatic or any infection, the VE estimates for bivalent vs. ≥3 monovalent (excluding the Singapore study) were 28.4% for all ages and 30.6% for seniors. For severe outcomes, the VE estimates for bivalent were comparable between all ages and ≥60 years regardless of the reference group, e.g., 72.9% and 72.0% for bivalent vs. unvaccinated, and 57.6% and 63.4% for bivalent vs. ≥2 monovalent. The VE of bivalent vs. ≥3 monovalent doses was also comparable between the two age profiles (62.1% after excluding the Singapore study vs. 60.7%). However, with most studies (8 out of 10) for all-age VE estimates conducted among seniors, this similarity could be partly attributed to the similarity in age profiles. Given that the senior age group is the most vulnerable to severe outcomes from COVID-19 [49], our findings suggest we should continue to prioritize the older subpopulation for coverage with bivalent vaccines.

Compared to three or more doses of a monovalent vaccine, the bivalent booster vaccines demonstrated superior relative efficacies against both infection (VE=28–42% depending on excluding or including the Singapore study) and severe disease (VE=68.5%) during the circulation of the BA.4, BA.5, and subsequent Omicron subvariants. This superiority can also be seen by calculating the relative efficacy of three or more doses of monovalent vs. unvaccinated. For example, in terms of severe disease, the relative efficacy of three or more doses of monovalent vs. unvaccinated is 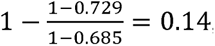, much lower than the relative efficacy of the bivalent booster vs. unvaccinated, 0.729.

Our study has several limitations. First, our analysis was constrained by the lack of long-term data, limiting our ability to assess the potential waning of VE over time. Some studies have shown the efficacy of the bivalent booster dose waned over time, similar to the monovalent booster doses [14,50], but the VE against severe outcomes generally wanes at a much slower rate than that against infection. Second, a few studies found prior infection could modify the efficacies of monovalent or bivalent boosters [37,50], but we were unable to stratify the VE estimates by prior infection status as most studies did not provide such information. Third, many studies included in this meta-analysis [21,22,29,32] did not specify the manufacturer or type of the bivalent booster and monovalent doses, introducing additional heterogeneity into and thus complicating the interpretation of our pooled results. This lack of granularity emphasizes the need for future studies to report VE estimates with greater specificity. Finally, we had to synthesize desired VE measures from multiple VE estimates in the same study, e.g., deriving the VE for the age group of 60 years or more from VE estimates for finer age groups. Such synthesis did not consider potential correlation among the finer estimates, which could result in under-evaluation of uncertainty in the synthesized VE estimate.

In conclusion, our study presents a systematic and up-to-date picture of the protective benefits of bivalent COVID-19 booster doses against Omicron subvariants, particularly against severe clinical outcomes. The SARS-CoV-2 pandemic continues to evolve with newly emerging variants that can better evade the current immunity landscape of the population but are not necessarily more pathogenic, e.g., HV.1 and JN.1 Omicron lineages have dominated previous strains in many places. While updated vaccines are rolling out at a slower pace, their strong protection against severe disease induced by the new strains has saved millions of lives and guards our daily life from interruption. On the other hand, as of September 2023, the coverage of bivalent vaccines in the U.S. remained below 50% even among people aged ≥60 years, according to the U.S. CDC. In September 2023, US CDC recommended vaccination with XBB-containing vaccines among persons aged ≥6 months [51]. In addition to developing more potent and convenient vaccines such as the flu-Covid combo vaccines, more resources should be directed to campaign the effectiveness and safety of existing bivalent vaccines, especially among the vulnerable subpopulations. Finally, we encourage future research on vaccine effectiveness assessment to provide more granular VE data and to monitor the long-term safety and effectiveness of bivalent/multivalent vaccines.

## Data Availability

The raw summary-level data were derived from publicly available papers cited in the reference. Extracted data and programming codes will be made available upon request by email to the corresponding author.

## Author Contributions

SSong, ZM, YY and IL conceived the study. SSong and ZM collected the data, SSong, ZM, ML, YM, SX, YH, SSarkar, AC and YY reviewed the data. SSong analyzed data under the supervision of YY, ZM and IL. SSong, ZM and YY drafted the manuscript. All authors contributed to interpretation of results. SSong, YY, ZM and IL finalized the manuscript.

