## Supplemental 1 for "A Systematic Review and Meta-Analysis on the Effectiveness of Bivalent mRNA Booster Vaccines against Omicron Variants"

**Appendix:**

**A.1 Supplementary Search Strategy: (resulting in 630 articles)**

**Search conducted time: November 8th, 2023.**

**Publication time: September 1st, 2022, to November 8th, 2023, if a date can be specified; 2022 to 2023 if the year is the most specific scope.**

Key words:

#1 (SARS-CoV-2) OR (COVID-19) OR (2019nCoV)

#2 bivalent booster

#3 (effectiveness) OR (efficacy)

#4 (test-negative) OR (case-control) OR (cohort study)

#5 Omicron

#6 (infected) OR (infection) OR (hospitalization) OR (hospital admission)

| **Database** | **Number of results** |
| --- | --- |
| **PubMed** | **18** |
| **Web of Science** | **6** |
| **Embase** | **29** |
| **Scopus** | **425** |
| **#1 AND #2 AND #3 AND #4 AND #5 AND #6 searched in all fields** | |

**Cochrane library only supports up to 5 search terms**

| **Database** | **Number of results** |
| --- | --- |
| **Cochrane Library** | **2** |
| **#1 AND #2 AND #3 AND #4 AND #5 searched in all fields** | |

**Preprint databases used different searching rules and limitations**

| **Database** | **Number of results** |
| --- | --- |
| **medRxiv** | **46** |
| **bioRxiv** | **4** |
| **for term "SARS-CoV-2 COVID-19 2019nCoV bivalent booster effectiveness test-negative case-control study Omicron infection hospitalization" and posted between "September 1st, 2022 and November 8th, 2023"** | |

**Google Scholar is a supplementary search source and can give many false positive results, so only the first ten pages of most relevant results will be screened.**

| **Google Scholar** | **Top 100 out of 3,290** |
| --- | --- |
| **((SARS-CoV-2) OR (COVID-19) OR (2019nCoV)) AND (bivalent booster) AND ((effectiveness) OR (efficacy)) AND ((test-negative) OR (case-control) OR (cohort study)) AND (Omicron) AND ((infected) OR (infection) OR (hospitalization)** | |

**A.2 PICOS Criteria Table:**

**PICOS Criteria**

| Participant | 1. Should be the general population excluding disease-specific or special condition cohorts (kidney cancer, pregnant women etc.). 2. We don’t restrict to any age group to the articles. |
| --- | --- |
| Intervention | 1. COVID-19 bivalent mRNA booster vaccination |
| Comparator | 1. Participants who did not receive bivalent booster, possibly including unvaccinated people (absolute VE), people received monovalent vaccines (relative VE). |
| Outcome | 1. VE=1-(adjusted odds/hazard ratio of Omicron infection and Omicron severe events). |
| Study Type | 1. Cohort study 2. Test-negative design study. 3. Or other observational study, e.g., Case-control study |
| Article Type | 1. Original research, instead of research plan, comments, review articles and so on. (Please denote if the review is relevant to bivalent VE in the exclusion reason column) 2. Must evaluate VE instead of other biological measurements, instead, evaluated neutralizing antibodies. |
| Note | 1. The paper published before August 31, 2022, might be not eligible. |

**A.3 Funnel Plots**

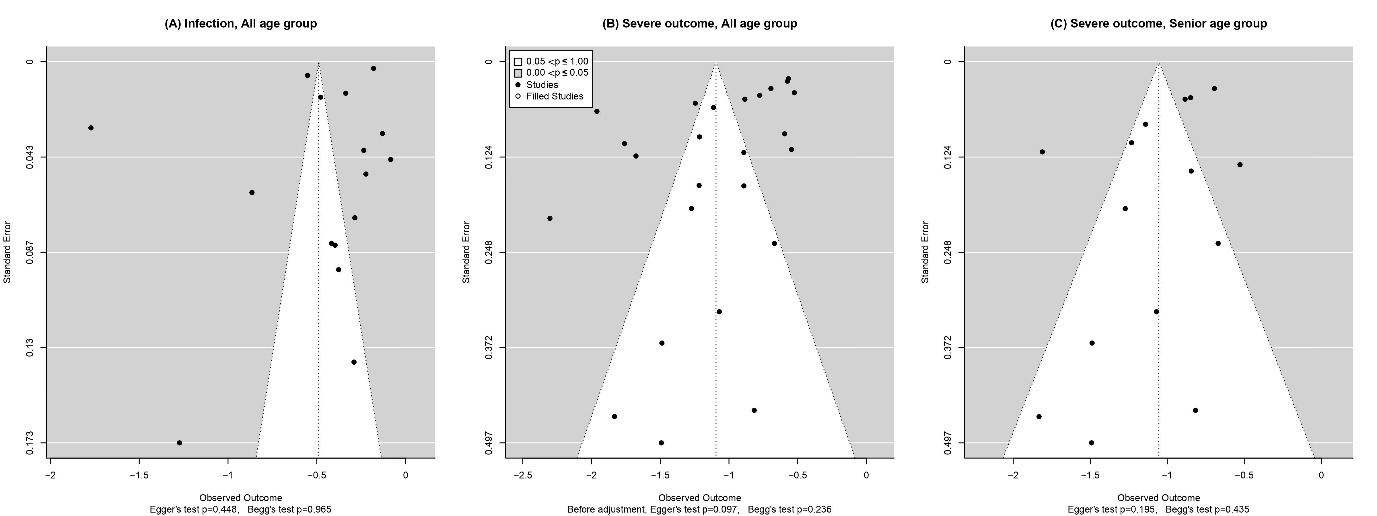

Figure S1. Funnel Plots for the pooled VE estimates.
